# Bioinformatic Analysis of Defective Viral Genomes in SARS-CoV-2 and Its Impact on Population Infection Characteristics

**DOI:** 10.1101/2023.10.05.23296580

**Authors:** Zhaobin Xu, Qingzhi Peng, Jian Song, Hongmei Zhang, Dongqing Wei, Jacques Demongeot

## Abstract

DVGs (Defective Viral Genomes) and SIP (Semi-Infectious Particle) are commonly present in RNA virus infections. In this study, we analyzed high-throughput sequencing data and found that DVGs or SIPs are also widely present in SARS-CoV-2. Comparison of SARS-CoV-2 with various DNA viruses revealed that the SARS-CoV-2 genome is more susceptible to damage and has greater sequencing sample heterogeneity. Variability analysis at the whole-genome sequencing depth showed a higher coefficient of variation for SARS-CoV-2, and DVG analysis indicated a high proportion of splicing sites, suggesting significant genome heterogeneity and implying that most virus particles assembled are enveloped with incomplete RNA sequences. We further analyzed the characteristics of different strains in terms of sequencing depth and DVG content differences and found that as the virus evolves, the proportion of intact genomes in virus particles increases, which can be significantly reflected in third-generation sequencing data, while the proportion of DVG gradually decreases. Specifically, the proportion of intact genome of Omicron was greater than that of Delta and Alpha strains. This can well explain why Omicron strain is more infectious than Delta and Alpha strains. We also speculate that this improvement in completeness is due to the enhancement of virus assembly ability, as the Omicron strain can quickly realize the binding of RNA and capsid protein, thereby shortening the exposure time of exposed virus RNA in the host environment and greatly reducing its degradation level. Finally, by using mathematical modeling, we simulated how DVG effects under different environmental factors affect the infection characteristics and evolution of the population. We can explain well why the severity of symptoms is closely related to the amount of virus invasion and why the same strain causes huge differences in population infection characteristics under different environmental conditions. Our study provides a new approach for future virus research and vaccine development.

## 1 Introduction

The SARS-CoV-2 virus poses a substantial and persistent threat to human life and health [1-3]. The study of this novel coronavirus has challenged traditional academic frameworks, such as the classical theory of herd immunity [4-6]. However, numerous puzzles remain unresolved, including the intriguing phenomenon of significant variations in population infection characteristics due to climate conditions and control strategies [7]. For example, during the extensive Omicron outbreak in China towards the end of 2022, individuals in northern regions exhibited a higher prevalence of severe symptoms, whereas the proportion of asymptomatic or mild cases was markedly elevated among those infected in southern regions when compared to their northern counterparts [8]. Another striking observation is the noticeably lower mortality rates in tropical areas like Singapore compared to Europe and North America, despite infections from identical or closely related viral strains [10]. These disparities cannot be simply attributed to differences in protective interventions, as such measures primarily influence infection rates rather than the severity of the disease, such as mortality. Deeper underlying factors may contribute to the observed variations in population infection characteristics. Leveraging bioinformatics analysis and mathematical simulations, our research uniquely attributes these distinctions to viral heterogeneity, specifically the presence of defective viral genomes (DVGs).

DVGs (defective viral genomes) are shorter and structurally different RNA molecules that arise during virus replication and are commonly present in RNA virus infections [11-13]. DVGs were first recognized by Preben Von Magnus in the late 1940s as incomplete influenza viruses capable of impeding the replication of the wild-type virus [14]. Over twenty years later, Alice Huang and David Baltimore introduced the term defective interfering (DI) particles, or DIPs, to describe viral particles that possess regular structural proteins but only a fraction of the viral genome. Furthermore, they specified that DIPs can only replicate in the presence of a helper virus and that they impede the intracellular replication of non-defective homologous virus [15]. Although the mechanism of DVG generation is not fully understood, it is generally believed that RNA polymerase plays a key role in their production [11,16-17]. RNA polymerase is an error-prone enzyme that lacks proofreading activity, which makes it prone to make mistakes during genome replication. These errors can lead to the synthesis of truncated or defective viral genomes that can serve as templates for the production of DVGs.

Previous studies analyzing high-throughput sequencing data found that DVGs or SIPs (semi-infectious particles) were widespread in RNA viruses [18-21]. The presence of DVGs can have both positive and negative effects on virus replication and pathogenesis. On one hand, DVGs can interfere with the replication of the full-length viral genome, leading to reduced viral titers and attenuated virulence. On the other hand, DVGs can activate the host immune response and enhance the production of antiviral cytokines, leading to increased viral clearance and improved immune protection [11,22-23]. Recent research has also been conducted on the role of DVGs in COVID-19 infection. One approach is to use DVGs as antiviral drugs, as studies have found that DVGs can significantly inhibit virus replication [24-25]. Sun Yan et al. also found significant differences in the content of DVGs among different cell types, and the proportion of DVGs changed significantly with the duration of infection [26]. They also noticed that the content of DVGs in asymptomatic individuals was significantly lower than that in symptomatic individuals. Although significant progress has been made in recent years regarding the study of viral heterogeneity, several key questions remain to be addressed. These include: (□) What are the mechanisms underlying the formation of viral heterogeneity, particularly defective viruses? (□) How does viral heterogeneity affect individual infection characteristics? (□) How does viral heterogeneity influence population infection characteristics and viral evolution?

To further elucidate the aforementioned issues, we have employed a comprehensive approach combining bioinformatics and mathematical modeling. Initially, we validated through viral second- and third-generation sequencing data that the generation of defective viruses in the SARS-CoV-2 virus is attributed to errors in RNA replication. Subsequently, utilizing mathematical models, we systematically investigated the impact of viral heterogeneity on individual and population infections. Our models provide a plausible explanation for the heightened transmissibility of late-stage SARS-CoV-2 variants and the effects of control measures and climate conditions on SARS-CoV-2 infection.

## 2 Materials and methods

### 2.1 Database

In our study, we employed NCBI virus database [27] to investigate the genomic information and sequence gaps of different strains. The NCBI Virus database provides comprehensive information on viral sequences and their characteristics, including genome sequences, SRA data, and detailed annotations. Therefore, we utilized this database to extract various sequencing results that contain genome and SRA information for different strains.

To access the SRA data and reference genome sequences of these strains, we used the SRA toolbox, a command-line tool developed by the NCBI. This tool enables users to download raw sequencing data in the form of SRA files, which can be converted into fastq files for further analysis. We combined these data with the corresponding reference genome sequences to obtain a more complete understanding of the genetic makeup of each strain.

### 2.2 Generation of genome depth spectrum and mining of DVGs

In the field of bioinformatics analysis, we have developed a novel algorithm that enables the rapid calculation of sequencing depth. This algorithm requires both the sequencing data and template gene sequences. With this approach, we can quickly calculate the short sequences in the sequencing data that match the template chain and identify breakpoints that occur during the sequencing process.

To apply this algorithm, first, we mapped the sequencing reads to the reference genome using a tool such as Bowtie or BWA. Then, we extracted the aligned reads that match the target template gene sequence from the output SAM/BAM file. Next, we calculated the sequencing depth at each position of the template gene sequence by counting the number of reads that map to that position. By comparing the sequencing depths across different regions of the template gene sequence, we can identify any regions with significantly higher or lower coverage than expected.

One key advantage of our algorithm is its speed. Compared with other methods, which may require significant computational time and resources, our algorithm provides rapid and efficient analysis. Additionally, our method can be used to detect breakpoints that occur during the sequencing process. Breakpoints are regions with gaps or deletions in the sequence that can cause errors in downstream analyses. By detecting these breakpoints, we can correct errors and improve the accuracy of subsequent analyses.

Overall, our algorithm offers a convenient and efficient method for calculating sequencing depth and identifying breakpoints in sequencing data. By accurately assessing sequencing depth, we can gain insight into the quality of the sequencing data and evaluate the performance of various sequencing platforms. Moreover, by detecting and correcting breakpoints, we can improve the accuracy and reliability of downstream analyses such as variant calling and de novo assembly.

### 2.3 Mathematical modeling of incomplete viral genome in individual infection

Unlike traditional models, we propose that viruses exhibit polymorphism, meaning that during the replication process, the complete RNA has a small probability of becoming partially deleted due to replication errors made by the polymerase. In practice, these deletions are continuous, but for simplicity in our study, we categorize them into three levels: intact RNA with no deletions (RNA_3), partially deleted RNA (RNA_2), and severely defective RNA (RNA_1). RNA_3 has the longest RNA chain and therefore has the longest replication cycle and the slowest replication rate, which we assume to be 1. However, there is a small probability (assumed to be 1%) that it may transform into RNA_2 during replication (k5 = 0.1, k6 = 0.9). RNA_2 has a shorter base length and a faster replication rate of 1.2. Similarly, there is a 1% probability of RNA_2 transforming into RNA_1 during replication (k3 = 0.12, k4 = 1.08). Additionally, we note that RNA_1 has lost the ability to generate functional viral proteins due to severe internal base deletions, while the translated protein activity of RNA_2 is partially impaired. Hence, we represent this weaker translation efficiency with k9 = 0.5. The translation efficiency of intact RNA_3 is 1 (k10 = 1). Although the likelihood of further deletions during virus replication is low, the shorter defective viral genomes have a faster replication rate and their proportion may significantly increase with each replication cycle. Another important feature of our model is the explicit representation of the interaction between antibodies and viruses, where processes 9-12 depict the binding reaction between antibodies and antigens, processes 13-16 represent the clearance of antigen-antibody complexes by the immune system, and processes 19-22 signify the regeneration of antibodies by antigen-antibody complexes.

The ordinary differential equations of the single-agent model are listed below. The definitions of variables are listed in Table 3. The parameters are defined in Table 1.

**Figure 1.**
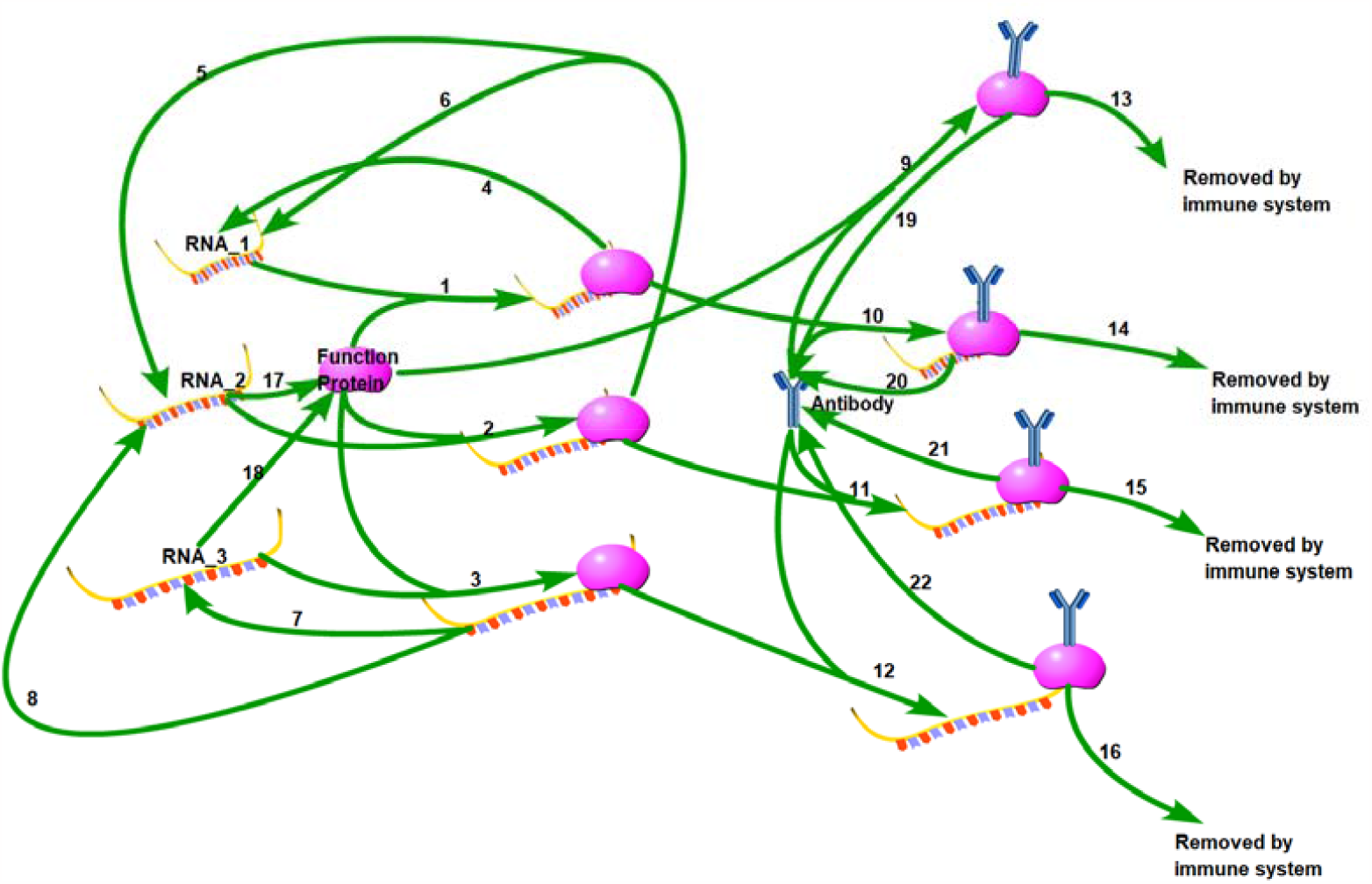
Biochemical scheme of SARS-CoV-2 life cycle. 22 reactions are illustrated in this diagram.

**Table 1.** Reaction index and the name of each reaction in our mathematical model.

| Reaction index | Reaction |
| --- | --- |
| 1 | $\text{RNA\_1} + \text{F} \xrightarrow{k_1} \text{RNA\_1-replication complex}$ |
| 2 | $\text{RNA\_2} + \text{F} \xrightarrow{k_1} \text{RNA\_2-replication complex}$ |
| 3 | $\text{RNA\_3} + \text{F} \xrightarrow{k_1} \text{RNA\_3-replication complex}$ |
| 4 | $\text{RNA\_1-replication complex} \xrightarrow{k_2} \text{RNA\_1}$ |
| 5 | $\text{RNA\_2-replication complex} \xrightarrow{k_3} \text{RNA\_2}$ |
| 6 | $\text{RNA\_2-replication complex} \xrightarrow{k_4} \text{RNA\_1}$ |
| 7 | RNA_3-replication complex $\xrightarrow{k_5}$ RNA_3 |
| 8 | RNA_3-replication complex $\xrightarrow{k_6}$ RNA_2 |
| 9 | F + Antibody $\xrightarrow{k_7}$ F-antibody complex |
| 10 | RNA_1-replication complex + Antibody $\xrightarrow{k_7}$ RNA_1-replication -Antibody complex |
| 11 | RNA_2-replication complex + Antibody $\xrightarrow{k_7}$ RNA_2-replication -Antibody complex |
| 12 | RNA_3-replication complex + Antibody $\xrightarrow{k_7}$ RNA_3-replication -Antibody complex |
| 13 | F-antibody complex $\xrightarrow{k_8}$ removed by immune system |
| 14 | RNA_1-replication -Antibody complex $\xrightarrow{k_8}$ removed by immune system |
| 15 | RNA_2-replication -Antibody complex $\xrightarrow{k_8}$ removed by immune system |
| 16 | RNA_3-replication -Antibody complex $\xrightarrow{k_8}$ removed by immune system |
| 17 | RNA_2 $\xrightarrow{k_9}$ F |
| 18 | RNA_3 $\xrightarrow{k_{10}}$ F |
| 19 | F-antibody complex $\xrightarrow{k_{11}}$ Antibody |
| 20 | RNA_1-replication -Antibody complex $\xrightarrow{k_{11}}$ Antibody |
| 21 | RNA_2-replication -Antibody complex $\xrightarrow{k_{11}}$ Antibody |
| 22 | RNA_3-replication -Antibody complex $\xrightarrow{k_{11}}$ Antibody |
| 23 | RNA_1 $\xrightarrow{k_{12}}$ Degradation |
| 24 | RNA_2 $\xrightarrow{k_{12}}$ Degradation |
| 25 | RNA_3 $\xrightarrow{k_{12}}$ Degradation |
| 26 | Antibody $\xrightarrow{k_{13}}$ Degradation |

**Table 2.** Estimates of the calibrated model parameters.

| Parameter Name | Description | Value | Range,refs |
| --- | --- | --- | --- |
| $k_1$ | Rate of Assembly constant between RNA and viral proteins | 5.0e-4 | [28 , 29] |
| $k_2$ | Replication rate of RNA_1(defective viral genome) based on RNA_1-replication complex | 1.5 | Assumed |
| $k_3$ | Replication rate of RNA_1(defective viral genome) based on RNA_2-replication complex | 0.12 | Assumed |
| $k_4$ | Replication rate of RNA_2(semi-infectious viral genome) based on RNA_2-replication complex | 1.08 | Assumed |
| $k_5$ | Replication rate of RNA_2(semi-infectious viral genome) based on RNA_3-replication complex | 0.1 | Assumed |
| $k_6$ | Replication rate of RNA_3(full viral genome)<br>based on RNA_3-replication complex | 0.9 | Assumed |
| $k_7$ | Antibody binding constant | 5.0e-4 | [28] |
| $k_8$ | Clearing rate of antibody-antigen complex | 0.5 | [28] |
| $k_9$ | Translation rate of viral functional proteins<br>based on semi-infectious RNA template | 0.5 | Assumed |
| $k_{10}$ | Translation rate of viral functional proteins<br>based on full-length RNA template | 1 | Assumed |
| $k_{11}$ | Activation constant of antigen-antibody<br>complex on antibody regeneration | 0.9 | [28] |
| $k_{12}$ | RNA degradation rate | 0.01 | [29] |
| $k_{13}$ | Antibody degradation rate | 0.01 | [28] |

**Table 3.** Time-dependent variables of the mathematical model characterizing the SARS-CoV-2 life cycle.

| Variables | Meaning | Initial value |
| --- | --- | --- |
| $x_1$ | Defective RNA | 0 |
| $x_2$ | Semi-infectious RNA | 0 |
| $x_3$ | Full length RNA | 1 |
| $x_4$ | Functional Protein | 0 |
| $x_5$ | RNA_1-replication complex | 0 |
| $x_6$ | RNA_2-replication complex | 0 |
| $x_7$ | RNA_3-replication complex | 0 |
| $x_8$ | Antibody | 1 |
| $x_9$ | F-protein-antibody complex | 0 |
| $x_{10}$ | RNA_1-replication -Antibody<br>complex | 0 |
| $x_{11}$ | RNA_2-replication -Antibody<br>complex | 0 |
| $x_{12}$ | RNA_3-replication -Antibody<br>complex | 0 |

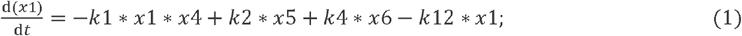

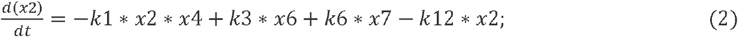

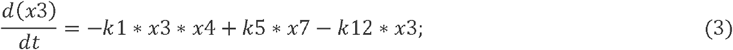

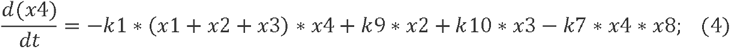

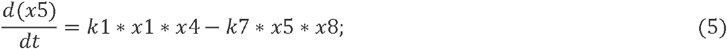

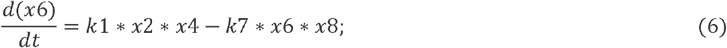

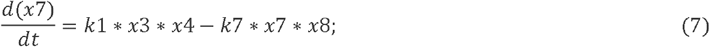

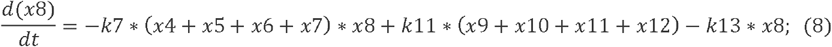

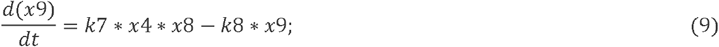

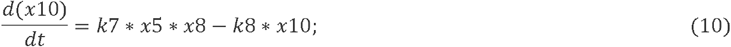

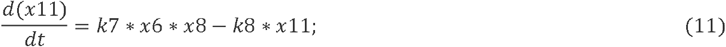

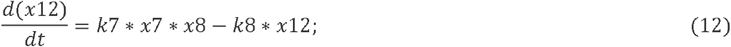

### 2.4 Mathematical modeling of incomplete viral genome in population infection

We proceed to incorporate the virus-antibody interaction within our agent-based model. In our previous study [30], we introduced a continuous Markov-chain model for simulating epidemics. This model considers a population consisting of N individuals, each exhibiting varying contact probabilities with others. The probability of infection is directly proportional to the contact probability, with the infection probability being equal to the contact probability itself. Notably, the contact probability between an individual and themselves is assigned a value of zero. Consequently, a matrix of size N × N is constructed, exhibiting the following properties:

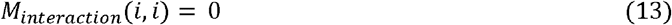

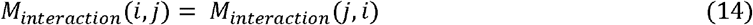

where *M*_*interaction*_(*i, j*) stands for the interaction possibility between individual *i* and individual *j*. An accurate contact matrix can be obtained by tracking the individual contact probability in a natural population group. For example, each person’s mobile phone can be recorded to obtain the population contact matrix within a particular time phase. The contact matrix is temporal and dynamic, which means it changes over time. However, it is difficult to obtain such accurate data at present. Therefore, the contact frequency is determined according to the relative distance between individuals, as shown in Equation (11).

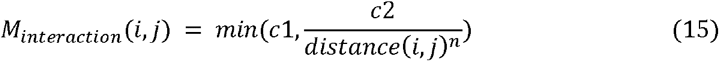

where *c*1 is the maximal contact possibility between agent *i* and agent *j*. In particular, the values of *c*1, *c*2, and *n* are preliminarily determined according to the initial reproduction constant *R*_*0*_ of the virus. According to the contact matrix, we can further determine the number of environmental invasive viruses received by a specific individual in a specific period of time.

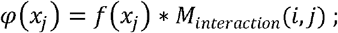

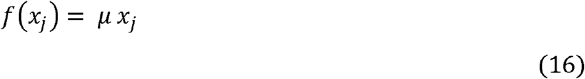

*φ*(*x*_*j*_) represents the inhaled virus of individual *i* from infected people *j. f* (*x*_*j*_) represents the overall released virus from infected person *j. f* (*x*_*j*_) is positive related to the virus loading *x*_*j*_ in corresponding infected agent with a correlation factor *μ*.

An extensive set of ordinary differential equations is further constructed. Assuming that the number of individuals in the population is N, each component is same to the single-agent model. A large set of equations are set as below:

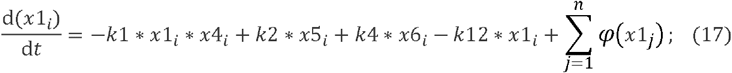

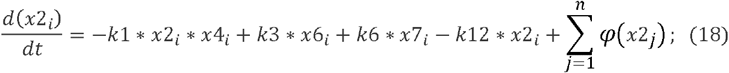

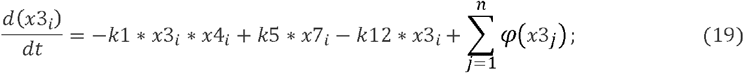

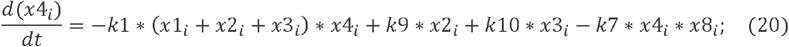

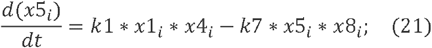

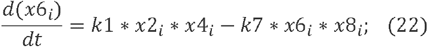

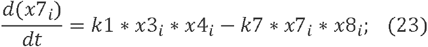

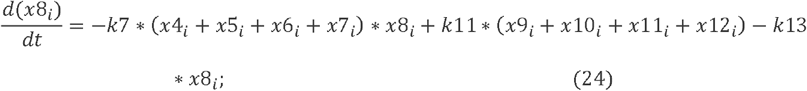

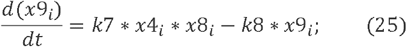

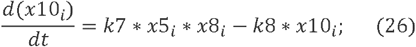

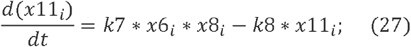

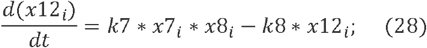

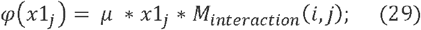

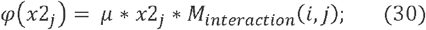

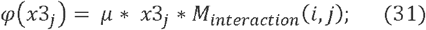

This system of equations contains 12*N variables, where the last term in equation (17-19) 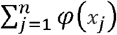 represents the overall number of viruses transmitted to this individual by other individuals in the whole population. We can generate random kinetic parameters that conform to the real population distribution through the first step of simulation. For such a large system of ordinary differential equations, we can solve it quickly by increasing the step size.

## 3 Results

### 3.1 Gene Sequencing Depth Profile Reveals Severe Heterogeneity in the SARS-CoV-2 Genome

The existence of genetic polymorphism in viral genomes, including incomplete genomes, has been recognized for a long time. However, there is currently a lack of systematic research on how defective viruses affect viral transmission and population infection characteristics. There are several possible causes of genomic defects in the SARS-CoV-2 virus, and we have summarized three highly plausible reasons for the formation of these defects. The first reason is the deletion in the UTR regions, which has been discussed in our previous articles. The second reason is genomic fragmentation, which may occur due to the action of host cell RNAase enzymes. The third reason is the generation of defective viruses, which will be defined in a new way in section 3.2, surpassing the understanding of many previous studies. Regardless of the cause of genomic heterogeneity in the virus, it can be reflected in sequencing depth profiles. Although fluctuations in sequencing depth at different loci may be influenced by various factors, a clear comparison between the genome depth profiles of DNA bacteriophages and SARS-CoV-2 reveals a greater degree of heterogeneity in the genome of the novel coronavirus. This genome heterogeneity is an important factor affecting viral population infection characteristics and viral evolutionary spread. We selected sequencing data from 10 types of DNA bacteriophages and 297 second-generation sequencing data of SARS-CoV-2, including 99 alpha strains, 93 delta strains, and 105 omicron strains. The results are shown in Figure 2. Figure 2a compares the sequencing depth of a bacteriophage with that of SARS-CoV-2. The coefficient of variation in gene sequencing depth can effectively measure the uniformity of the genome. As evident from Figure 2b, the coefficient of variation in gene sequencing depth of SARS-CoV-2 is significantly greater than that of DNA bacteriophages, indicating a highly pronounced heterogeneity in the genome of the novel coronavirus. The completeness and uniformity of the genome in omicron virus sequencing are notably superior to delta and alpha strains.

**Figure 2a.**
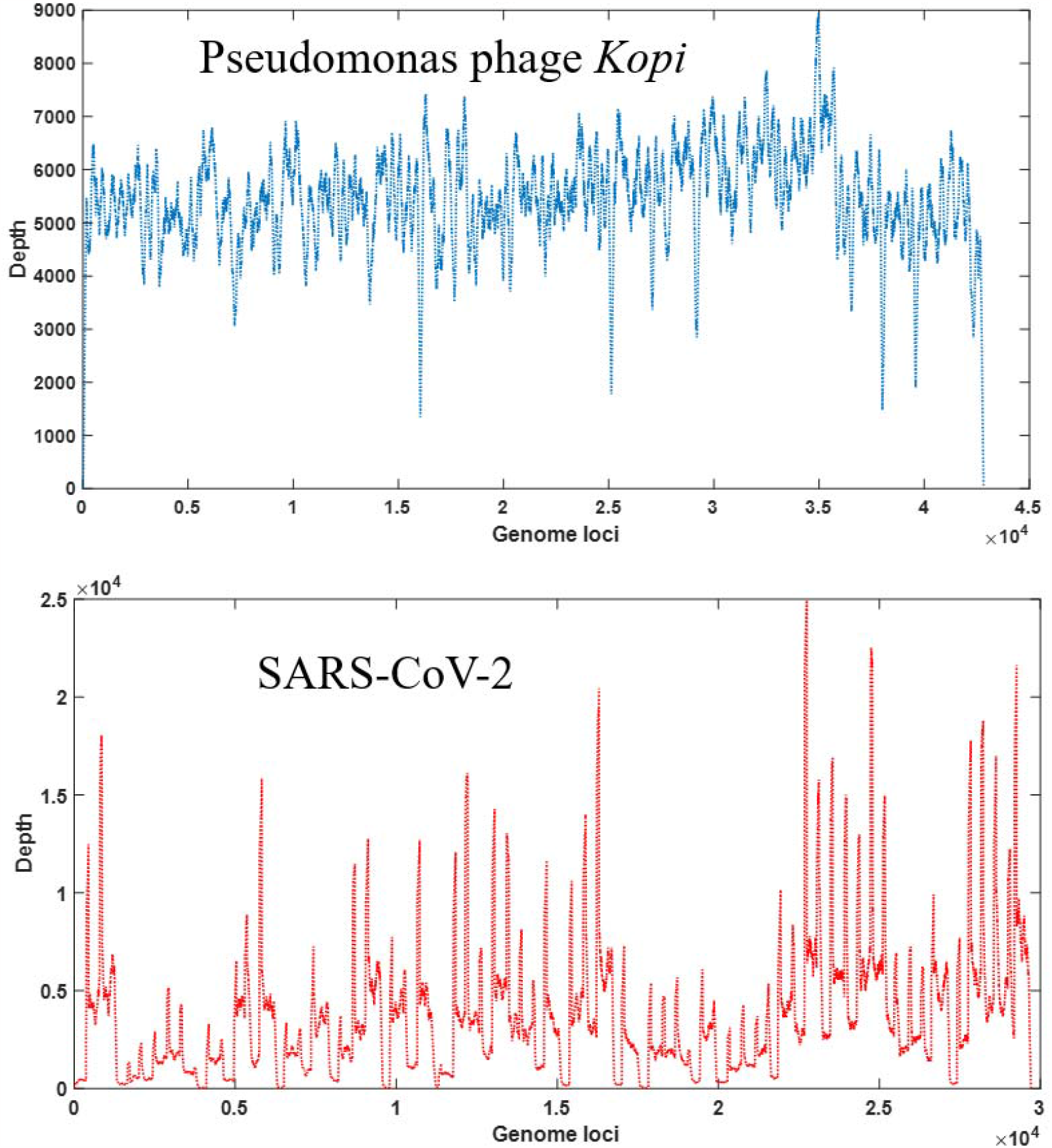
Depth coverage of DNA bacteriophage (SRR16248203) and SARS-CoV-2 (SRR23175982).

**Figure 2b.**
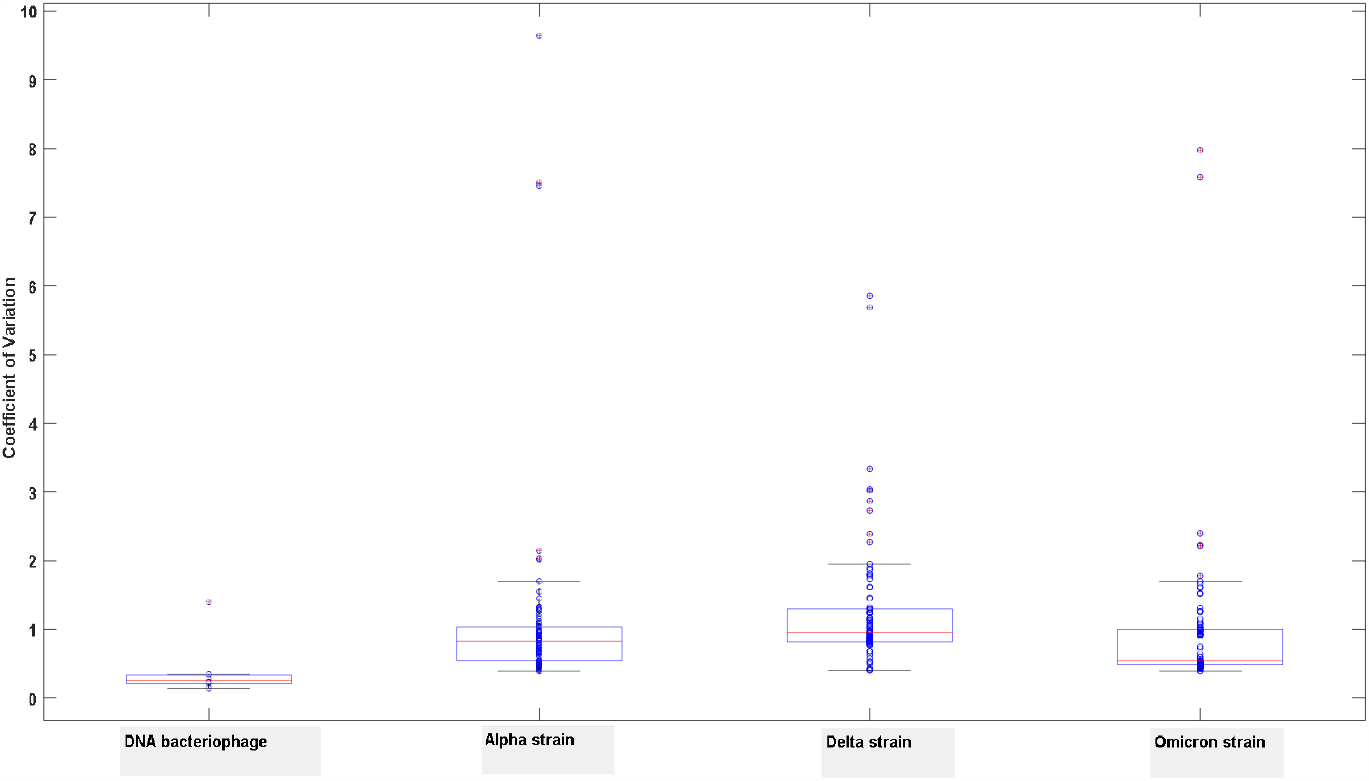
Uniformity of NGS data among DNA bacteriophage and three major strains of sars-cov-2

### 3.2 Comparison of Defective Viruses in Different SARS-CoV-2 Strains

Here, we provide a new definition for defective viruses. Traditionally, defective viruses are generally defined as viral particles with partial genomic deletions. When these deletions are significant, the encoded functional proteins are significantly affected and often unable to complete replication, hence referred to as defective viruses. There is no essential difference between defective viruses and semi-infectious particles (SIP). When the deleted fragment is relatively short, the virus’s replication function is affected but not entirely lost, thus referred to as SIP. Here, we propose a new definition for defective viruses, which includes any alteration to the original viral genome apart from point mutations that affects its normal biological function. This greatly expands the scope of defective viruses. Based on this definition, we can classify defective viruses into two categories. The first category consists of the traditionally studied defective viruses, where there is significant internal genomic deletion. The second category comprises defective viruses with internal duplicated segments. These two cases can be represented by the pattern diagram shown in Figure 3a.

**Figure 3a.**
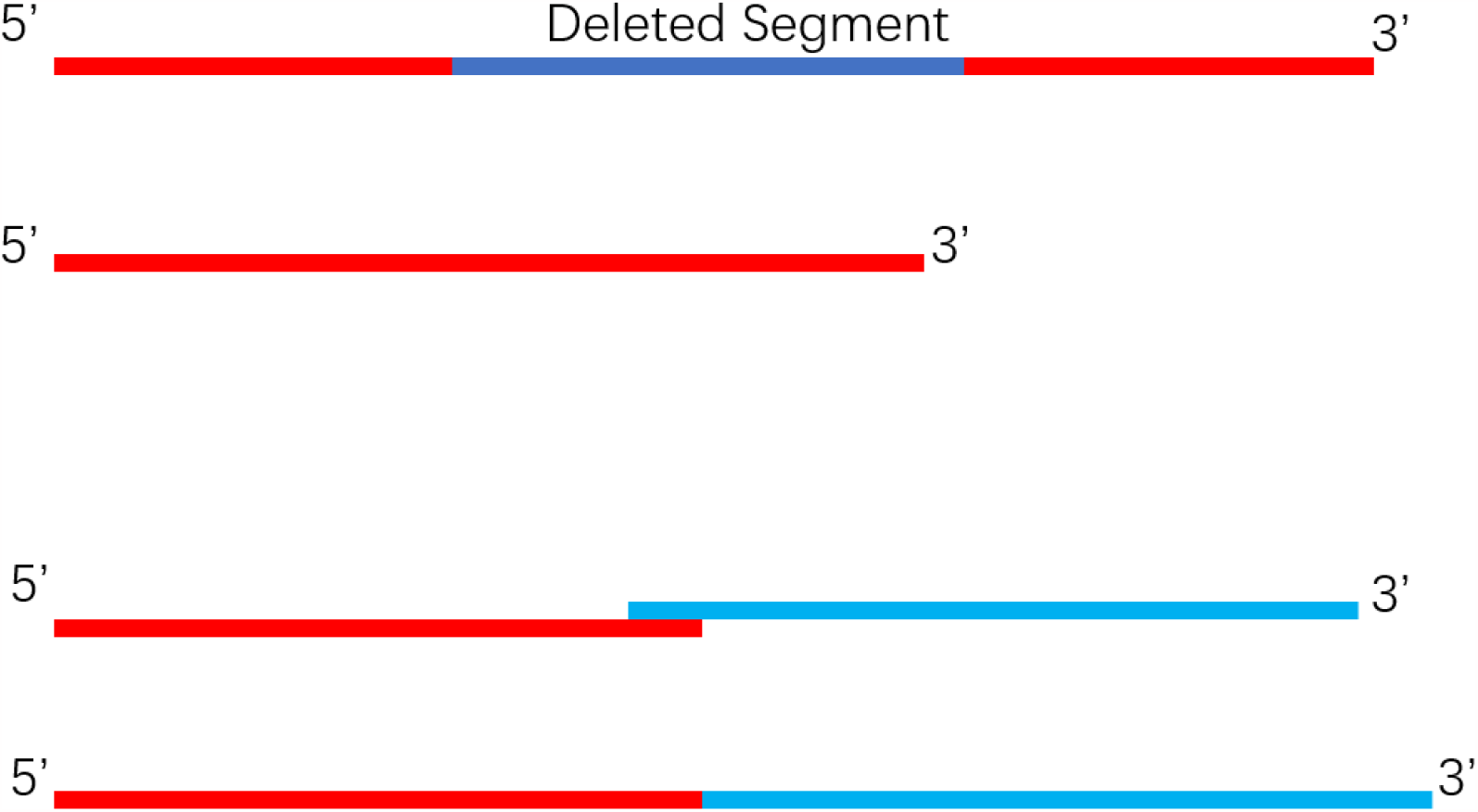
A diagram of two types of defective viruses

**Figure 3b.**
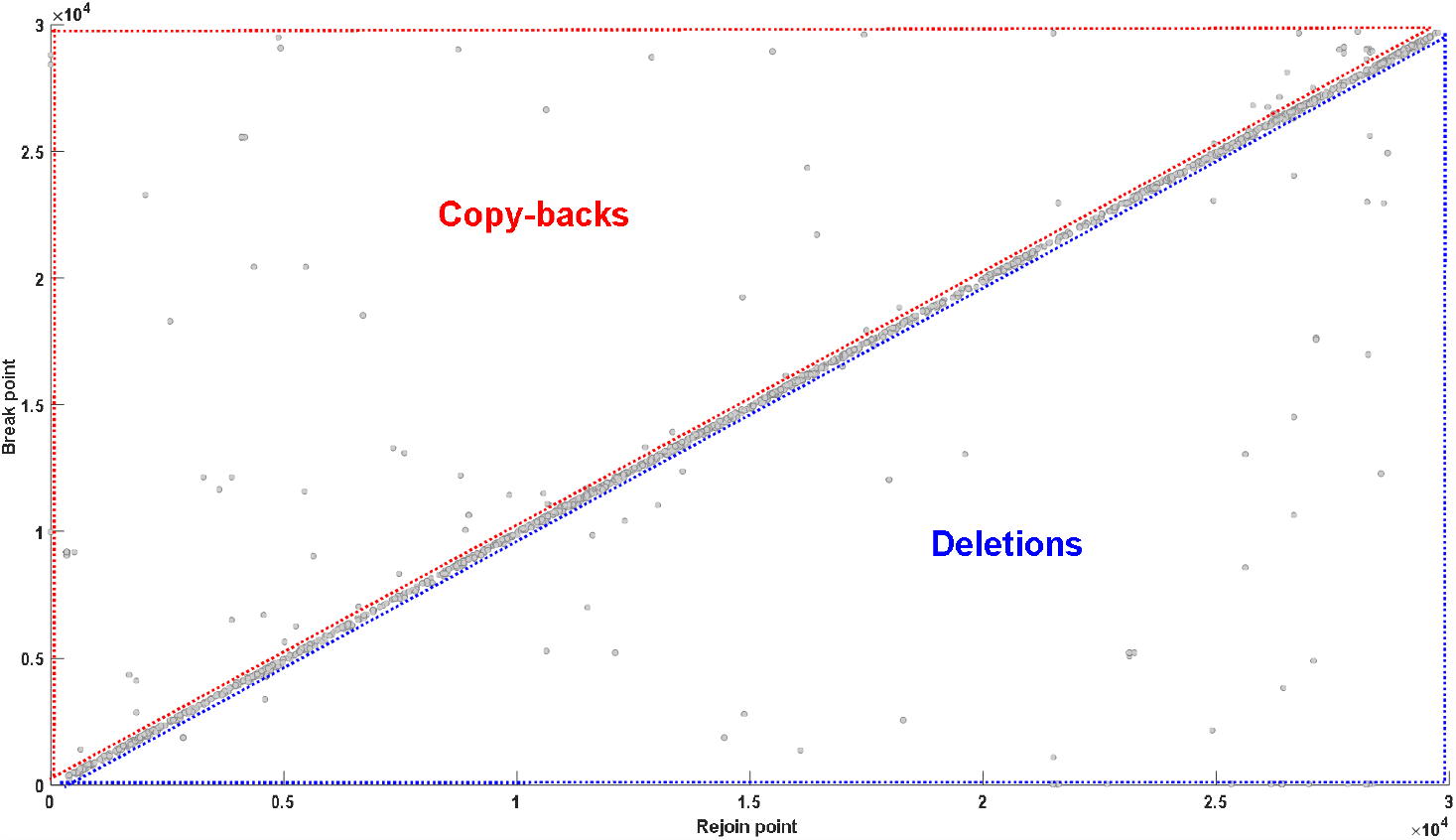
Analysis for junction distribution of DVGs identified in NSG data

**Figure 3c.**
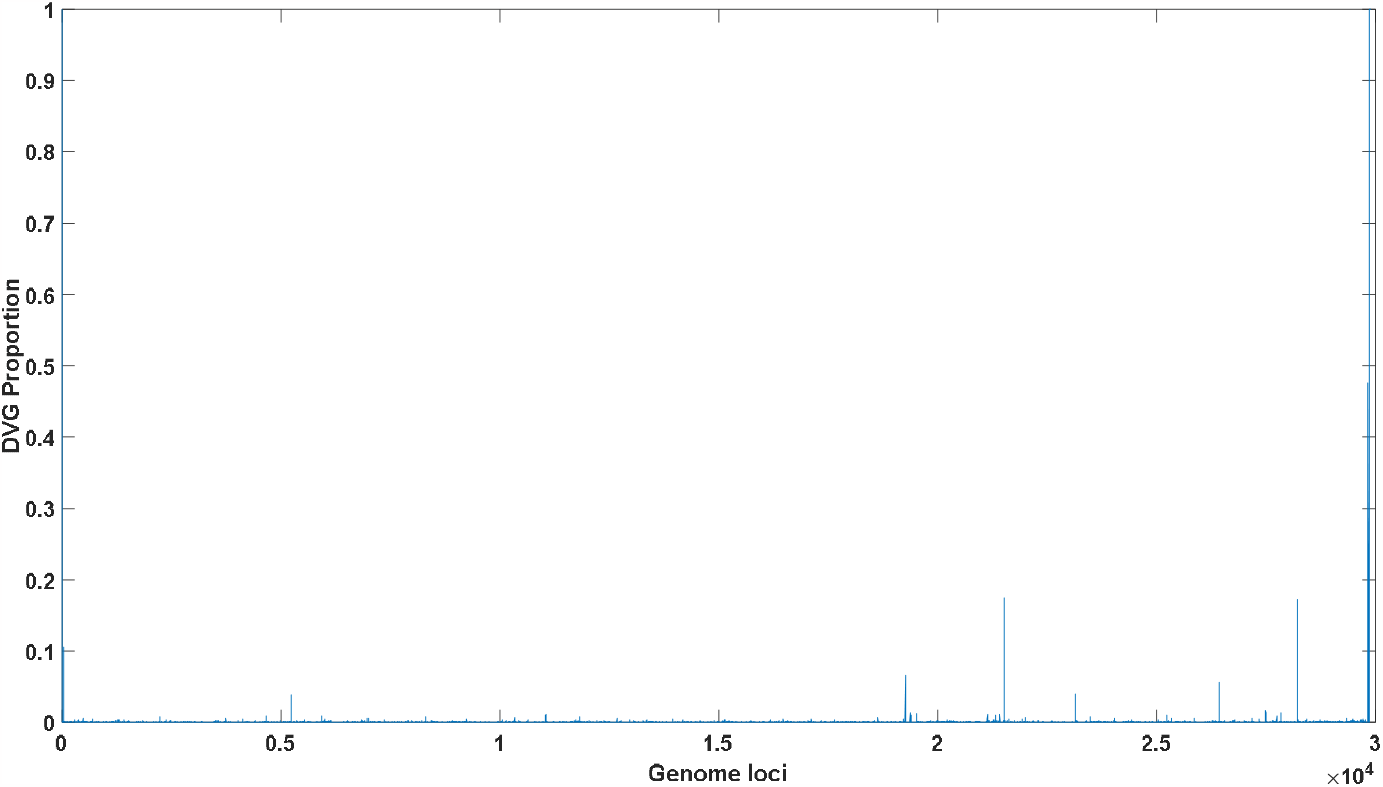
Analysis of DVGs transformation probability in each genome loci

**Figure 3d.**
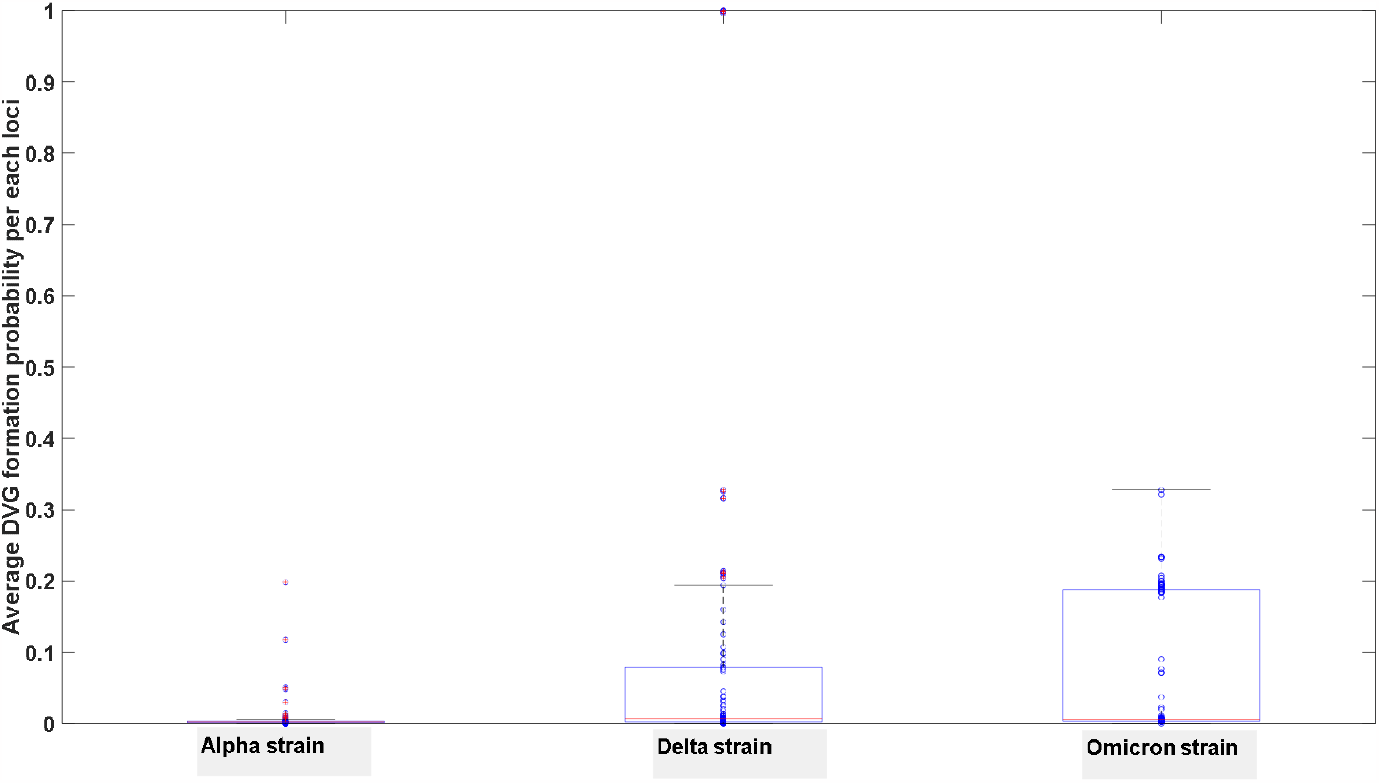
Comparison of DVGs overall transformation probability among three major strains of SARS-CoV-2

For the first case, the sequence number of the new 3’ end generated by deletion on the left is obviously smaller than the sequence number of the new 5’ end on the right, indicating a deletion has occurred. In the second case, the sequence has been elongated, which is also an important cause of defective virus formation, known as copy-backs. We set a threshold of 5nt, where deletions or insertions of duplicated segments exceeding 5 nt are considered typical defective viruses. The two-dimensional site map of defective viruses is depicted in Figure 3b, from which we can observe the specific sites where deletions occur or where replication duplication takes place. The probability of deletion or erroneous insertion at each site is represented in Figure 3c. By using Figure 3c, we can calculate the average occurrence of insertions or deletions across the entire genome. We analyzed the data from 297 SARS-CoV-2 second-generation sequencing samples, including 105 Omicron, 93 Delta, and 97 Alpha strains, and the results are shown in Figure 3d. Figure 3d represents the average probability of each nucleotide position experiencing a defective-type mutation. These mutations are not point mutations but may involve breakage or copy-backs. Figure 3d demonstrates that all strains can give rise to defective viruses, and the occurrence of defective virus mutations in the Omicron strain did not significantly decrease. Although the proportion of intact genomes in the Omicron strain is significantly higher than that in the Delta and Alpha strains, as observed in Figure 4c later, the proportion of defective viruses is only closely related to the time of infection and is not strongly associated with the specific infecting strain.

**Figure 4a.**
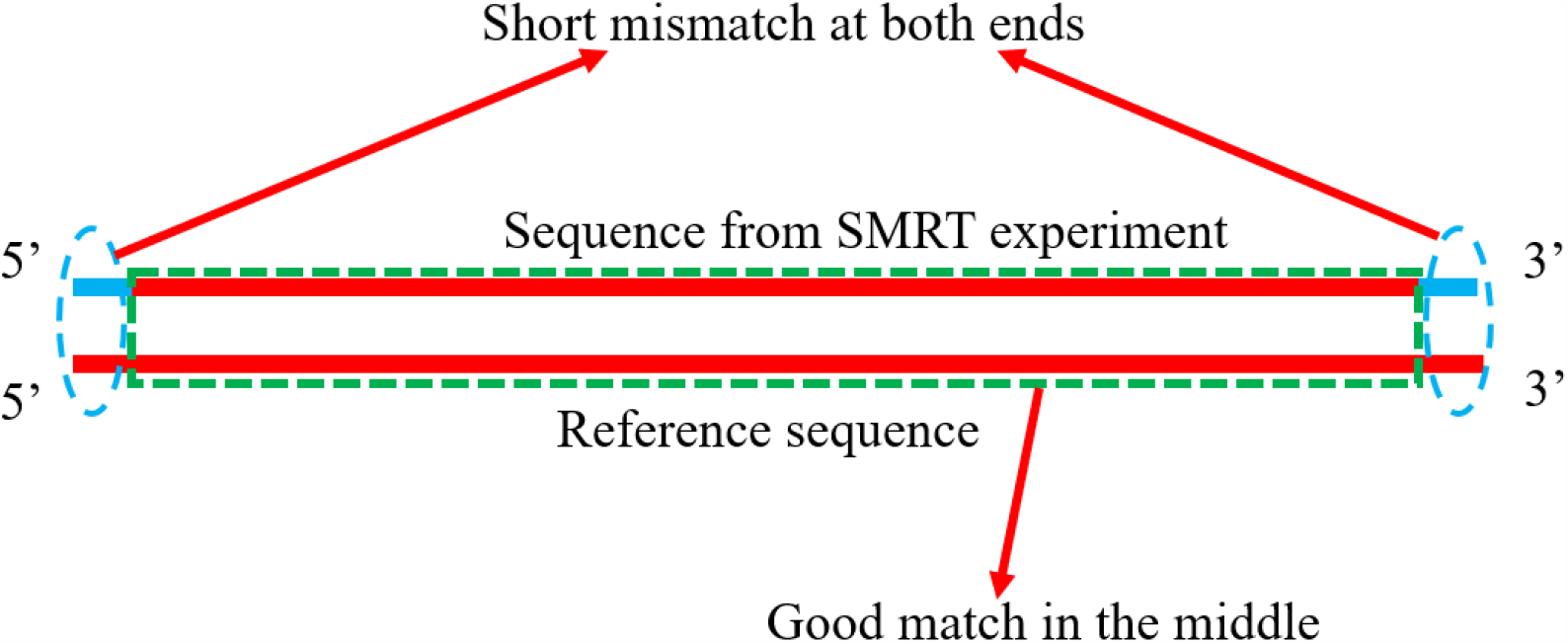
A diagram of replication error caused by SARS-CoV-2 RNA polymerase

**Figure 4b.**
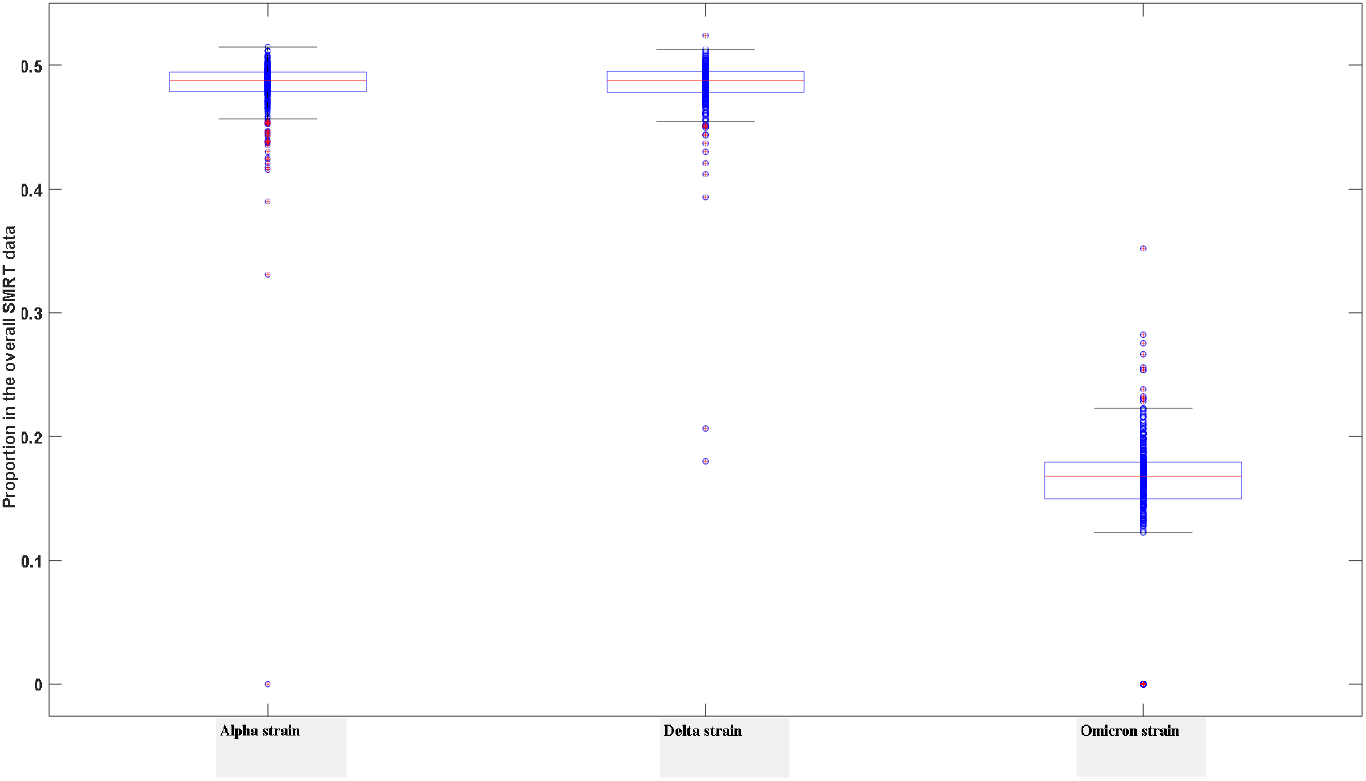
Proportion of end mutations in the overall SMRT data in three major SARS-CoV-2 strains.

**Figure 4c.**
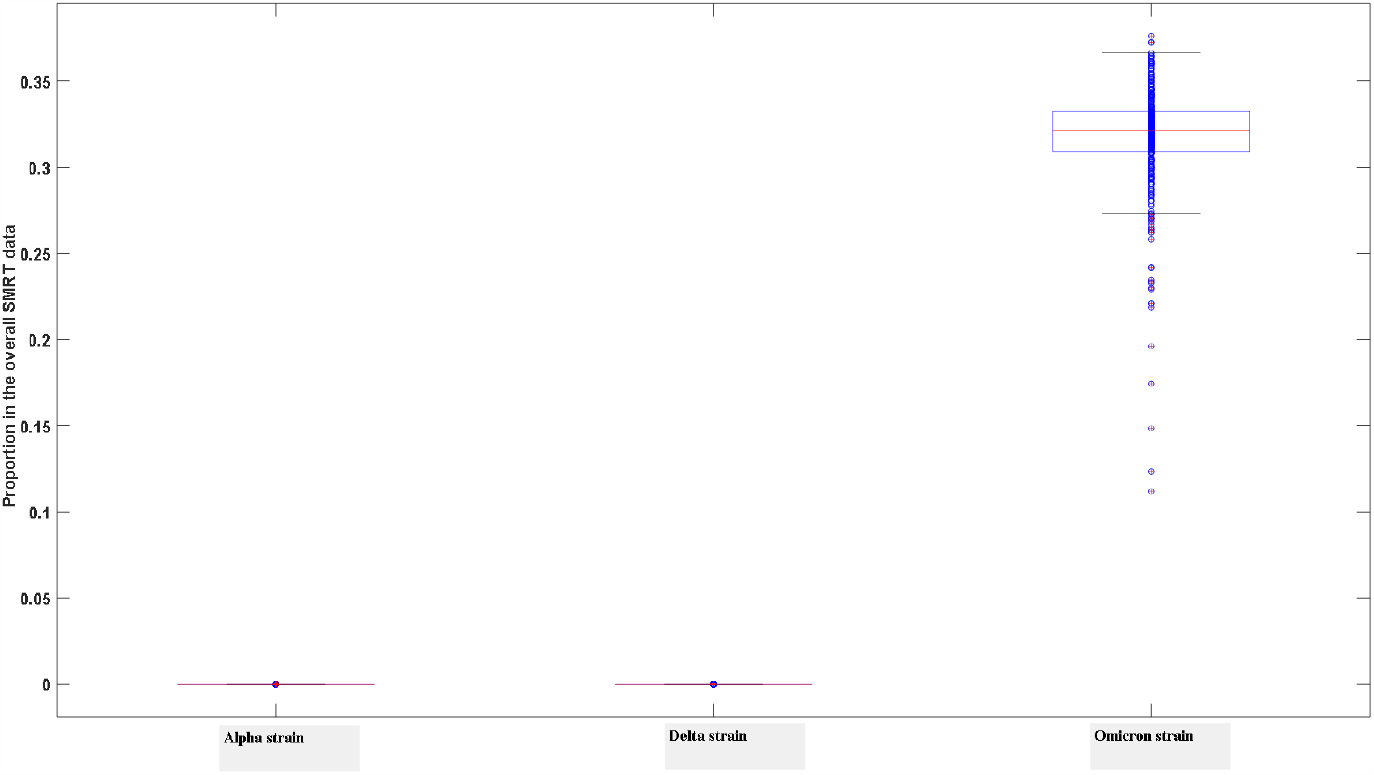
Proportion of full-matched segments in the overall SMRT data in three major SARS-CoV-2 strains.

### 3.3 The mechanisms of defective virus generation revealed by third-generation sequencing data lie in the replication errors of RNA polymerases

Currently, there is no consensus on the causes of defective virus generation. One widely accepted theory is that replication errors occur in RNA polymerases. However, another possibility is that viral RNA undergoes cutting by host RNAases and recombination mediated by RNA ligases. In this study, we utilized third-generation sequencing data to demonstrate that this alternative mechanism either does not exist or is extremely rare. If the second mechanism were operative, the recombinant viral genome would inevitably contain fragments of host RNA due to potential ligation of host transcript fragments during the RNA ligation process. Initially, we employed bioinformatics methods to filter out RNA sequences that do not match the viral reference genome in both positive and negative strands, as well as their reverse complements. Consequently, these sequences could only originate from exogenous RNA or accumulated replication errors. The sequences are provided in the supplementary materials. By comparing them with the human genome and transcriptome, we found no evidence that any RNA fragment originated from the human transcriptome. Therefore, we reject the second mechanism and attribute defective virus generation to replication errors in RNA polymerases.

By comparing the alpha, delta, and omicron data, we discovered that for the alpha and delta strains, the third-generation sequencing data showed hardly any sequences that fully matched the reference genome. In contrast, a considerable proportion of sequences from the omicron strain exhibited complete matches with the reference genome. For the alpha and delta strains, the non-matching regions of long sequences were predominantly located at the sequence ends, while the middle regions displayed high degrees of matching. This phenomenon is not due to exceptionally high fidelity of the RNA polymerase in the omicron strain but rather reflects two important mechanisms. Firstly, the replication process of the SARS-CoV-2 RNA polymerase exhibits an extremely high error rate, which may surpass our previous understanding. During the early stages of replication, a template sequence with a high mismatch rate is synthesized. Subsequently, the proofreading ability of proteins such as Nsp corrects the erroneous bases. The long and short sequences generated by third-generation sequencing effectively capture these details. In the process of reverse transcription, these RNA polymerases are shed, resulting in severe base mutations at both ends of the DNA sequence, while the middle region tends to exhibit a high degree of matching. This mechanism is illustrated in Figure 4a. The second mechanism is that the packaging capacity of the omicron strain significantly exceeds that of the delta and alpha strains, meaning that the number of viral particles is much higher in the former. RNA encapsulated by the capsid proteins often consists of relatively intact RNA sequences with 5’ and 3’ structures, at least after completion of replication. Such RNA sequences do not carry RNA polymerases. Therefore, we observed a large number of long sequence segments from the omicron strain that fully matched the reference genome. These fully matching long sequence segments originate from RNA sequences enclosed by capsid proteins. However, for the alpha and delta strains, due to their significantly lower assembly capabilities compared to the omicron strain, complete chains are prone to degradation. Hence, the proportion of intact RNA fragments in the third-generation sequencing data that match the reference sequence is extremely low, making it difficult to detect long sequences that fully match the reference sequence. Moreover, for the omicron strain, a significant number of long sequences with end mutations were also observed, indicating that the reliability of RNA polymerase replication has not significantly changed compared to previous strains. This is illustrated in Figure 4b, where all mutant strains exhibit a considerable proportion of long sequences with end mutations. Although the Ct values of Omicron in nucleic acid detection (quantitative PCR) may be slightly higher than those of other strains, such as delta, its genome integrity is significantly greater. This can also explain why Omicron exhibits exceptionally high infectivity. The proportions of sequences that fully match the reference genome in third-generation sequencing data for different strains are shown in Figure 4c. From Figure 4c, it can be observed that the omicron strain contains a large number of complete genome sequences, which is significantly higher than that of early strains such as delta and alpha.

### 3.4 The Impact of Defective Viruses on Individual Infection

While experimental studies have already confirmed the disruptive role of defective viruses in viral replication, there is currently a lack of systematic research on the impact of defective viruses on individual infection. The presence of defective viruses plays a crucial role in viral evolution and clearance within the host organism.

#### 3.4.1 Defective virus can prevent severe infections

Traditional research on host-virus interactions often focuses on the host’s immune function. However, in reality, besides activating the host’s immune system, viral replication errors leading to the production of a large number of defective viruses are also an important factor in their eventual clearance by the immune system. Without the occurrence of replication errors caused by viral polymerases, infected individuals may trigger more severe immune responses and exhibit more severe clinical symptoms.

**Figure 5a.**
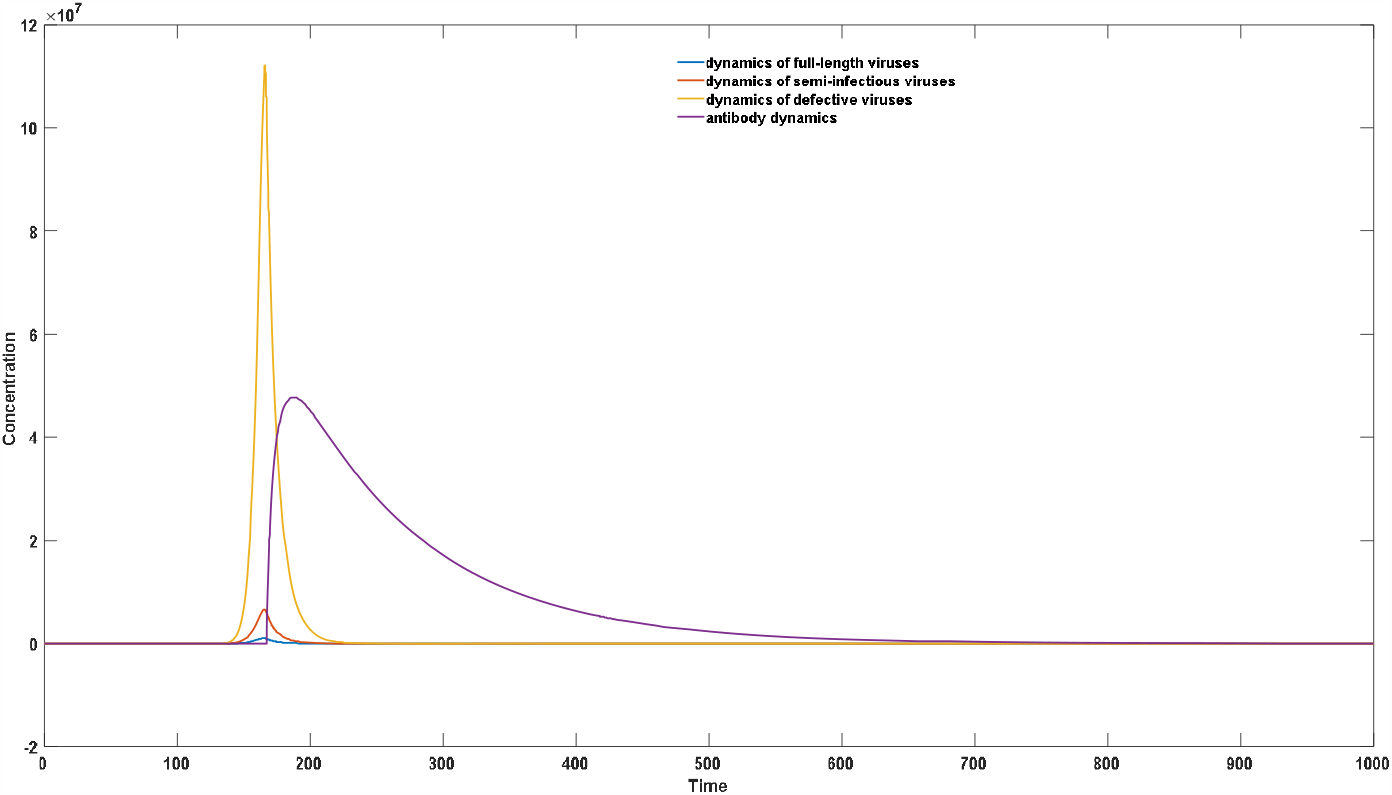
Virus-antibody dynamics after semi-infectious virus infection

**Figure 5b.**
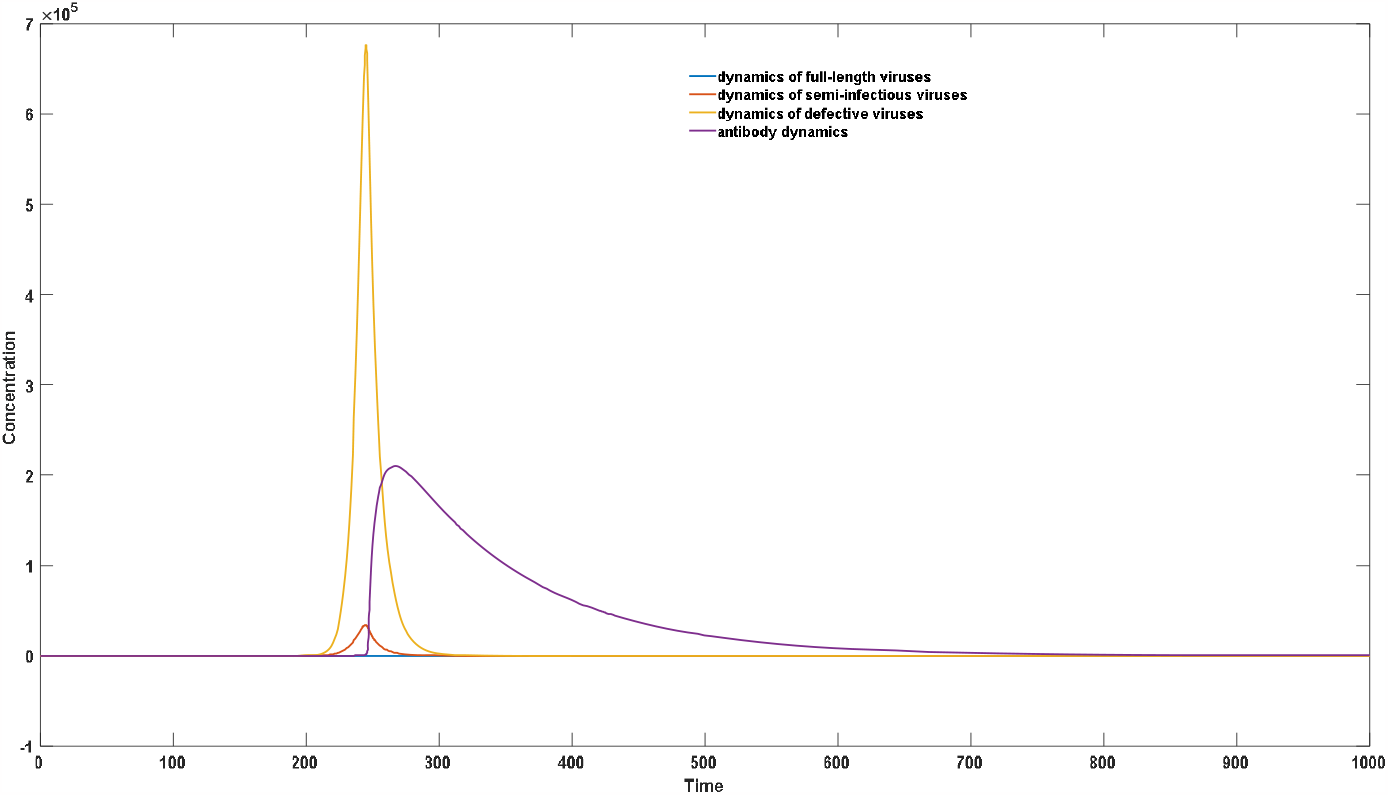
Virus-antibody dynamics after full-length virus infection (replication error = 10%)

**Figure 5c.**
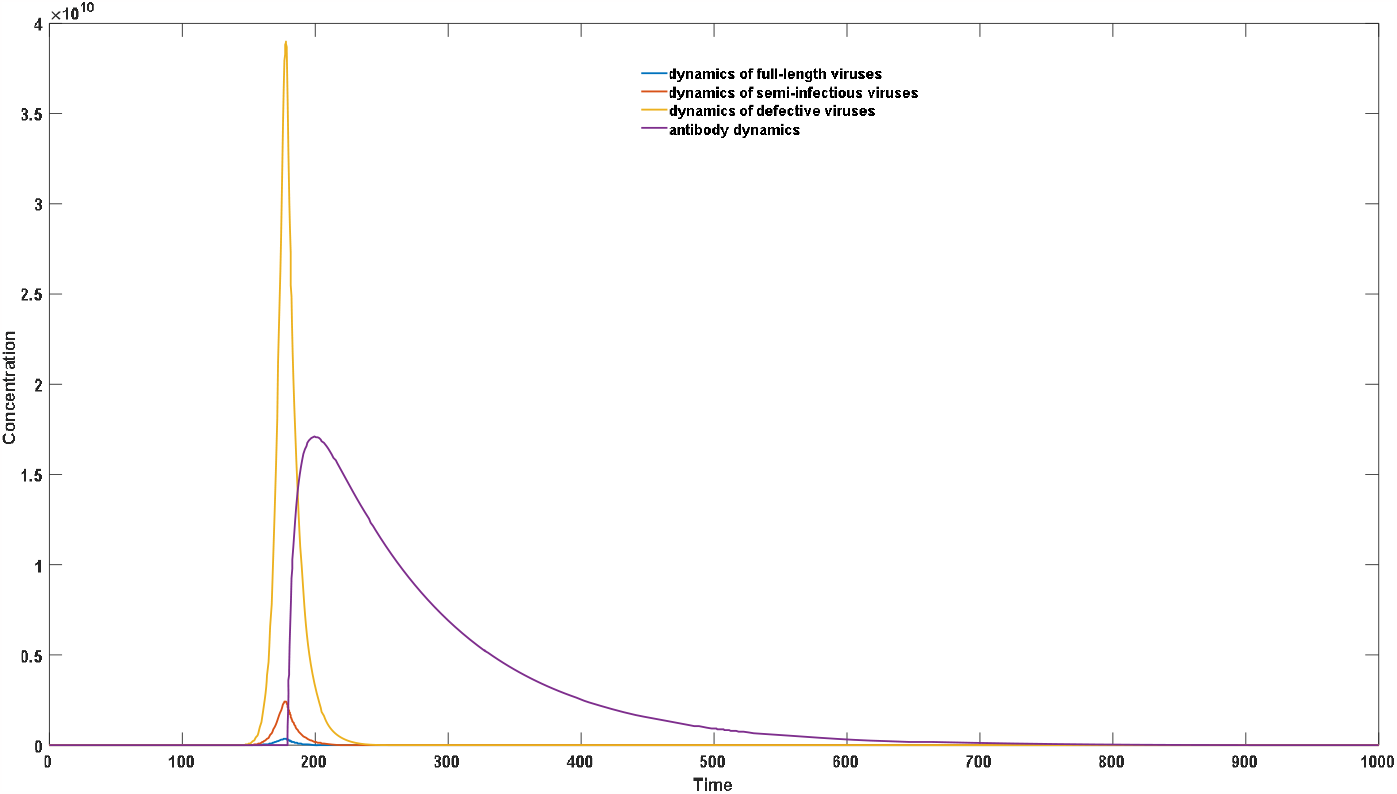
Virus-antibody dynamics after full-length virus infection (replication error = 5%)

Figure 5 visually illustrates that the severity of infections caused by semi-infectious viruses is significantly lower than that caused by full-length viruses. Additionally, a comparison between Figure 5b and 5c indicates that the generation of replication errors plays a crucial role in reducing the severity of infection. When the replication error rate decreases, i.e., when the probability of generating defective viruses during each round of replication decreases, both the peak viral load and peak antibody concentration significantly increase. This will manifest as more severe clinical symptoms.

#### 3.4.2 Generation of defective viruses can reduce the infectivity of individuals in the late stage of infection

As reported in experimental studies, the proportion of defective viruses significantly increases with the progression of the infection cycle [26]. Simultaneously, there is a significant decrease in the proportion and absolute concentration of intact RNA viruses. This phenomenon explains why patients exhibit strong infectivity during the incubation period and early stages of infection, while their infectivity rapidly declines in the later stage of infection. This can be depicted in Figure 6.

**Figure 6.**
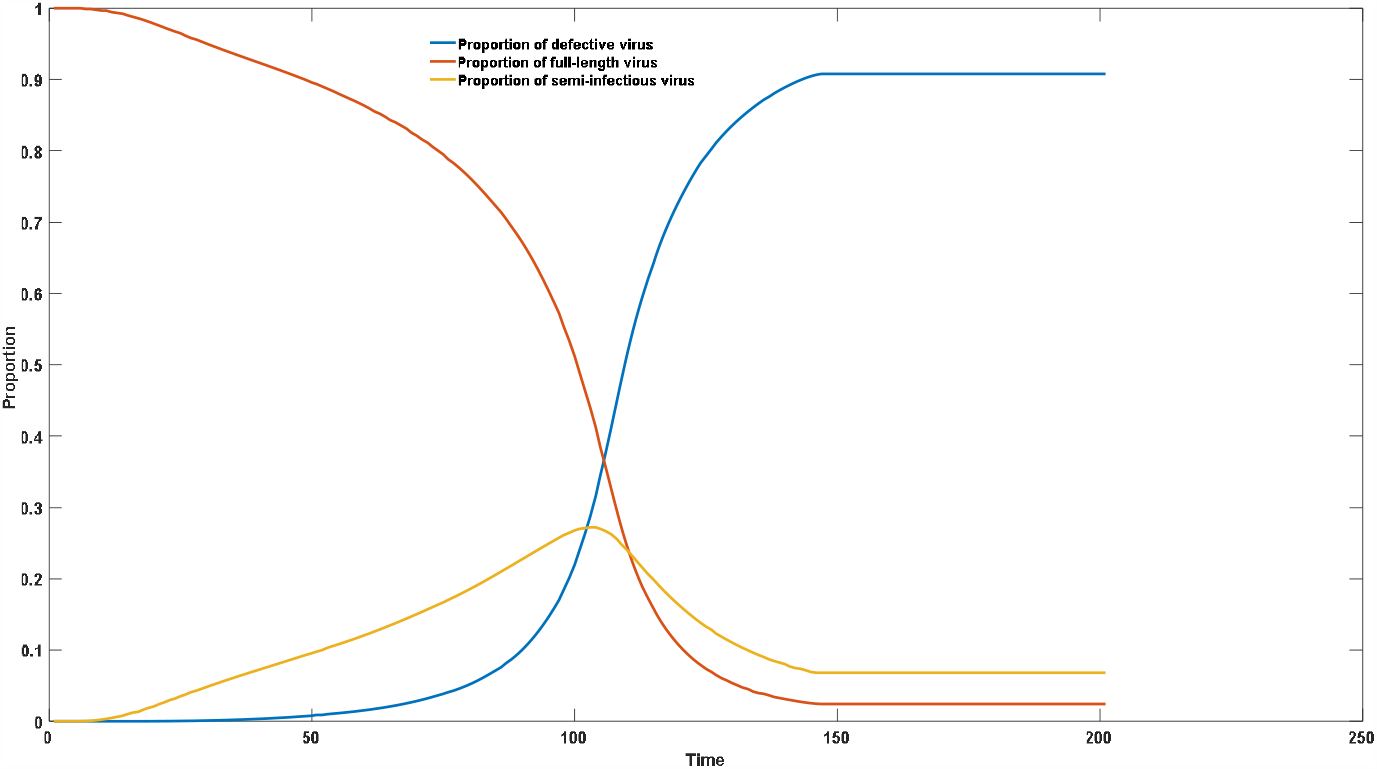
Dynamics of virus composition after infection

#### 3.4.3 The host antibody level exerts selective pressure on intact viruses, screening out full-length viruses to ensure that the virus does not succumb to rapid decay caused by replication errors

Without considering the interaction between host and virus, it is easy to reach the conclusion that the virus will rapidly evolve towards lower virulence. For example, let’s consider a simple scenario where there is a probability p of replication errors and the generation of defective viruses during each round of replication. After N rounds of replication, the proportion of intact viruses would decrease to (1-p)^N^. Regardless of how small the value of p is, when N is large enough, the proportion of intact viruses will gradually decline until they eventually disappear. However, the actual process of viral evolution is not as straightforward. Take respiratory viruses such as SARS-CoV-2 and influenza virus for instance. Despite the existence of a typical mechanism for generating defective viruses, their virulence can remain stable for a considerable period of time, thanks to the host’s selective pressure against highly virulent viruses. This may sound somewhat paradoxical, but the screening effect of antibodies on highly virulent intact viruses is indeed significant. In the absence or presence of extremely low levels of antibodies, all self-replicating viral particles, including semi-infectious viruses, can invade the host and proliferate. However, when the host’s antibody level reaches a certain threshold, weakly replicating genomic segments of defective viruses fail to effectively proliferate. Conversely, when highly replicating intact viruses invade, the antibody level is insufficient to completely inhibit their replication, allowing the viruses to proliferate effectively. Hosts often experience a substantial increase in antibody levels after initial infection; however, over time, these levels gradually decline. The antibody level first decreases to a threshold that permits replication of intact viral particles, and then declines to a threshold that allows replication of defective viruses. Through this mechanism, the host tends to preferentially select intact, highly active viral particles during secondary infection, rather than those with defects. This specific pattern is illustrated in Figure 7. It should be noted that not all viruses are fortunate enough to undergo this process. When the value of p is large, the population’s evolution can still rapidly progress irreversibly towards loss of virulence. In our previous research articles [31], we suggested that the rapid disappearance of SARS and MERS may be attributed to this phenomenon, as truncation of the two UTRs is a common way to generate defective viruses with weak replication activity.

**Figure 7a.**
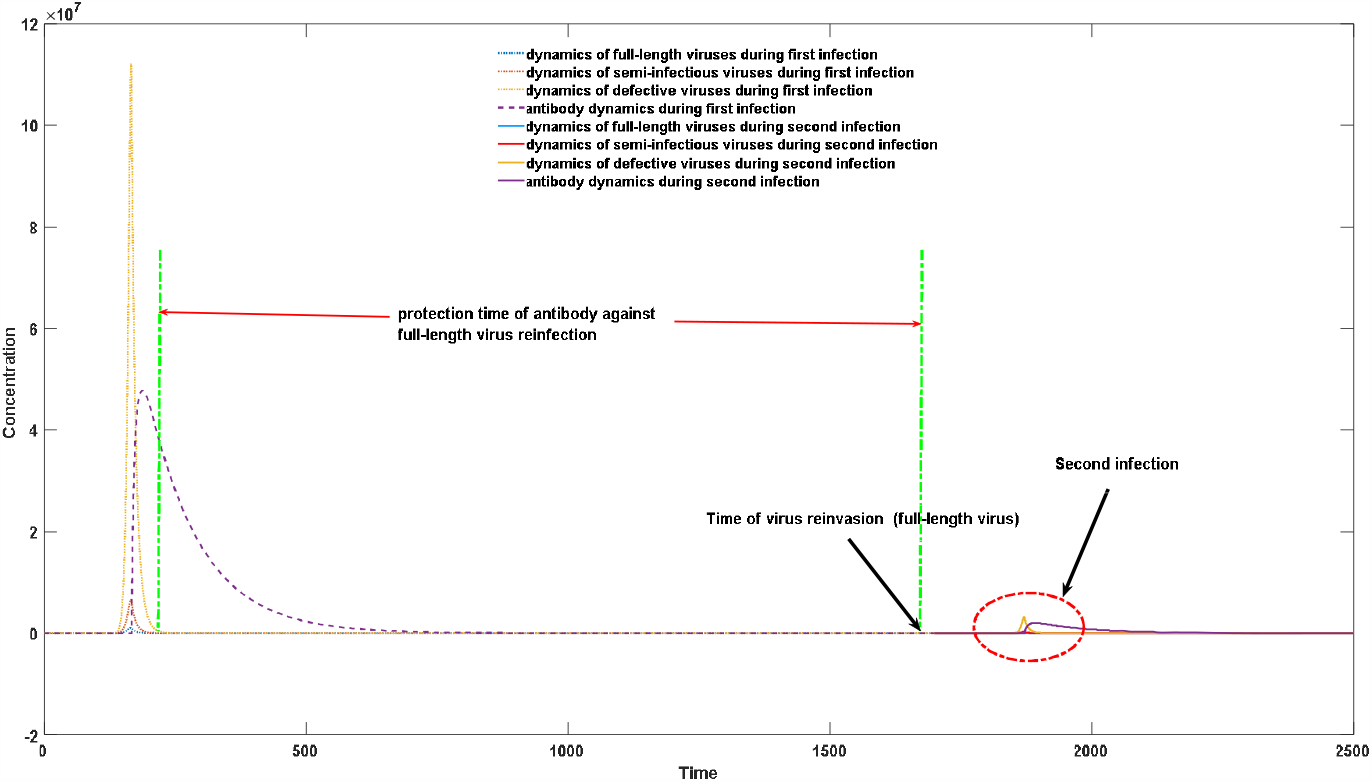
protection time against reinfection with full-length virus

**Figure 7b.**
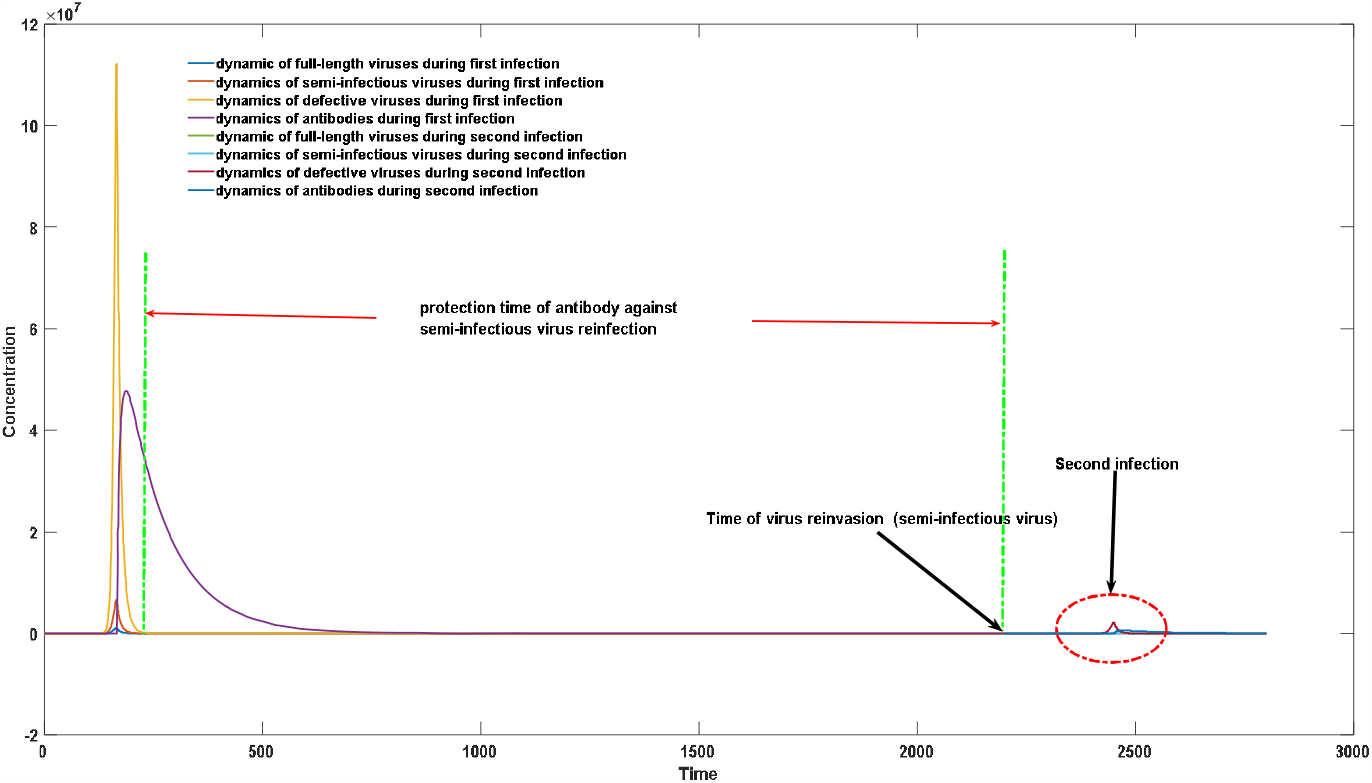
protection time against reinfection with semi-infectious virus

From Figure 7, it can be observed that the increased antibody levels following initial infection have varying durations of protection against different viruses. The duration of protection against semi-infectious viruses is significantly longer than that against full-length viruses. This gives rise to a phenomenon where, during the time interval between viral invasion at 7a and the invasion of semi-infectious viruses at 7b, the host is susceptible to full-length viruses but immune to semi-infectious viruses. This selection pressure favors the persistence and evolution of full-length viruses.

### 3.5 How Defective Viruses Impact Population Infection: A Study on the Infectious Characteristics

Extensive research has been conducted on defective viruses from the perspectives of experimental and bioinformatics studies. However, there is a significant lack of research on how defective viruses influence the infection characteristics of host populations. Defective viruses may have profound effects on the infection characteristics of populations, which could greatly challenge our traditional understanding of infectious disease transmission processes. Many infection characteristics of populations are associated with the presence of defective viruses.

To illustrate this phenomenon, let’s consider a simple example: low temperature and dry climate favor virus transmission, and many respiratory infections tend to have major outbreaks during the winter season. Moreover, these lower temperature conditions often lead to more severe symptoms. For instance, during the late 2022 Omicron outbreak stage, patients in northern China generally experienced more severe symptoms compared to asymptomatic or mild cases prevalent in southern regions. Protective measures also significantly impact the symptoms experienced by infected individuals. When society adopts strict control measures or individuals take stringent protective measures, not only can the likelihood of infection be effectively reduced, but also the severity of infection can be significantly diminished. All these commonly observed phenomena are closely related to the presence of defective viruses.

Before employing mathematical models to systematically simulate this process, we can provide a macro-level explanation. Both experimental and theoretical studies indicate that the peak viral load after infection has little correlation with the initial invading viral dosage. Instead, it is determined by the virus’s replicative ability and the host’s immune response. This implies that protective measures only reduce the probability of infection but not the severity of infection. However, for SARS-CoV-2 infections, the actual situation suggests that reducing the initial invading viral dosage helps mitigate the symptoms experienced by infected individuals. This has been prominently demonstrated in statistical studies on the effectiveness of mask protection. The only explanation lies in the heterogeneity of viral infections, meaning that despite belonging to the same strain of the novel coronavirus, there are significant genetic differences in their genomes.

To illustrate this, let’s consider a simple example: if the proportion of intact viral particles within the total viral population is 1%, then when a host inhales a virus, they are inhaling a proportion of intact viral particles equal to 1%. However, when they inhale N particles, the proportion of intact viral particles reduces to 1-0.99^N^. Additionally, there are numerous semi-infectious particles and completely non-functional defective viruses present in the air. Completely non-functional defective viruses, lacking autonomous replicative ability, do not cause infections. Conversely, due to their competitive binding with functional proteins produced by normal viruses, mixed infections often lead to an overall decrease in viral replicative capacity, resulting in milder clinical symptoms. Defective viruses with partial functionality and minor deletions exhibit weaker replicative abilities compared to normal viruses, leading to lower peak viral loads and weaker antibody elevation levels, resulting in less severe infection symptoms. Complete viruses that can infect individuals will cause severe infections. Therefore, if individuals are infected with semi-infectious particles, they will exhibit milder infection symptoms, and such infections remain transmissible, resulting in subsequent infections that also manifest less severe clinical symptoms. Mathematical models can better simulate and explain this macro-level phenomenon. Here, we have utilized and further developed our agent-based model based on antibody kinetics, and specific explanations of this model can be found in the Methods section.

By adjusting μ, representing the number of environmental viruses inhaled by each individual in the population, we can modulate different protective measures, as well as the influence of environmental conditions and temperature on infection. A higher μ value signifies lower control measures or colder environments, which are more prone to an increase in the number of environmental viruses inhaled by individuals. Conversely, a lower μ value signifies strict control measures or hot and humid external environments. From Figure 8, it can be observed that effective control measures and humid climate conditions significantly reduce the peak antibody levels and viral loads in the population, corresponding to milder infection symptoms.

**Figure 8.**
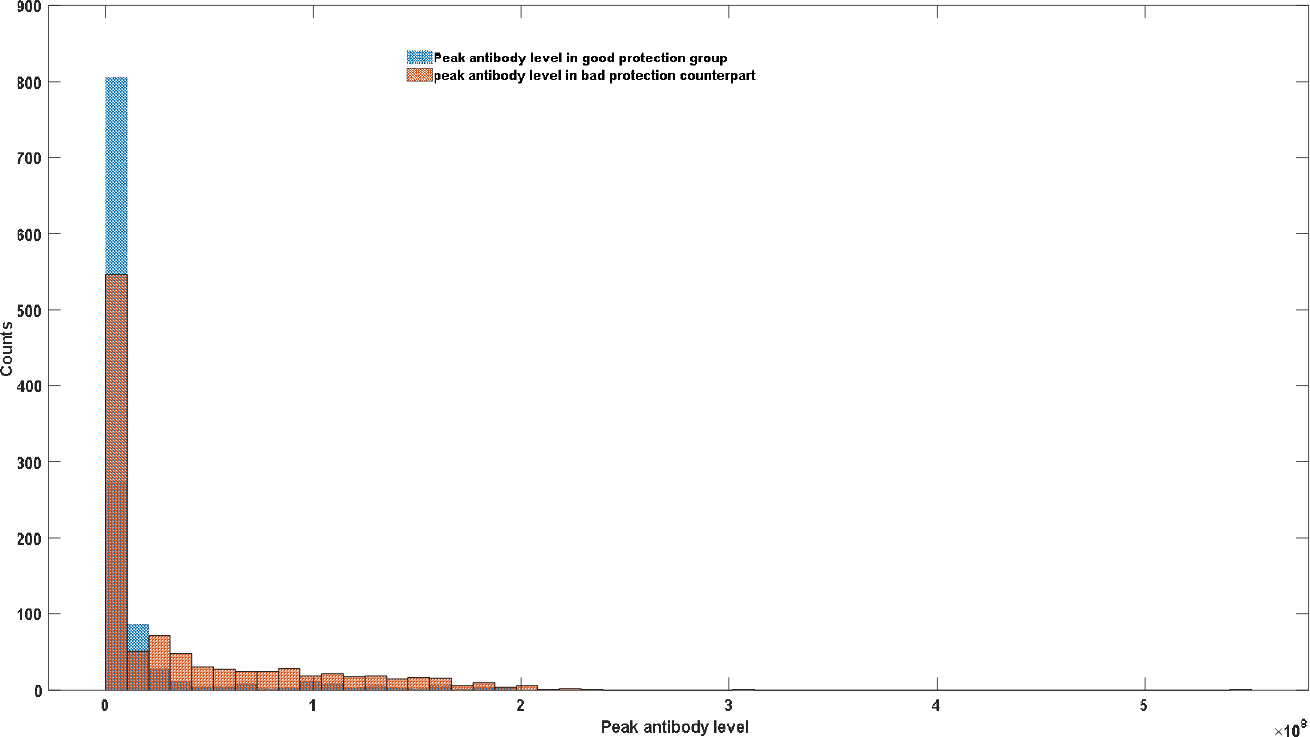
Effects of protection measure on the infection severity at population level.

## 4 Discussion

As our understanding of defective interfering viral genomes (DVGs) deepens, we are gradually realizing that viral infection is no longer a homogeneous and simple process. Even if the genetic types of invading viruses are completely identical, their completeness may vary greatly. Traditional DVGs are considered to be viruses with replication defects because their genomes have severe deletions. However, sequencing results show that the fragmentation and recombination of viral genomes is largely a random process, and this can lead to significant differences in the completeness of viral genomes.

This random loss inevitably produces a large number of semi-infectious viruses with low degrees of deletion. These semi-infectious viruses can have a profound impact on population infection characteristics. Most traditional virus genome studies focus on how the full genome characteristics of viruses affect pathogenicity. However, the existence of DVGs and semi-infectious viruses suggests that the completeness of viral genomes is also an essential factor in determining viral pathogenicity.

Focusing only on sequencing data for mutation mining can cause many problems, including the inability to accurately assess the pathogenicity or mortality rate of a strain, which can lead to major errors in public policy. A vivid example is the contradiction and confusion among infectious disease experts during the outbreak of COVID-19 in China, especially after the emergence of the Omicron strain. Mortality data in different regions showed significant deviations, even for the same virus subtype, with death rates in tropical areas such as Singapore significantly lower than those in other developed countries such as North America. In early December 2022, China lifted its epidemic prevention policy, and before that, the mortality rate caused by the Omicron strain in China was very low. However, when the epidemic prevention policy was lifted, Omicron caused a large number of deaths in China, conservatively estimated at more than 5 million. Most importantly, unlike before, most infected individuals in the north of China exhibited high fever symptoms, rather than mostly asymptomatic infections. However, this situation was not so severe in southern China, where the proportion of asymptomatic infections was significantly higher than in the north.

Our theory of viral heterogeneity can explain this phenomenon well. Our simulation results also show that, under the premise of the existence of the theory of viral heterogeneity, there is a significant correlation between population infection symptoms and the total amount of virus inhaled. Therefore, effective prevention and control measures and a hot and humid environment can help alleviate overall infection symptoms, but are not conducive to the survival and evolution of intact virus particles. The disappearance of many viruses, such as SARS in 2003, and the Wuhan SARS-CoV-2 in early 2020, were not due to excellent prevention and control measures leading to the complete disruption of the virus transmission chain and thus the disappearance of the virus. Excellent prevention and control measures can reduce the total amount of virus inhaled, and this decrease in inhaled virus amounts accelerates the evolution of the virus towards lower toxicity and more severe deletion of semi-infectious particles. Therefore, what we first eliminated was high-toxicity, intact viruses rather than the virus itself.

A vivid example is when the Wuhan epidemic ended completely in 2020, more than 300 positive infections were still found in the Wuhan region through mass nucleic acid testing conducted in June 2020. However, these positive infections could not stably transmit and replicate within the host body [32]. This suggests that it is necessary to pay attention to the completeness of viral genomes and the production of DVGs when designing public health policies and implementing prevention and control measures.

The COVID-19 epidemic is almost over, but the possibility of future similar infectious diseases is still high. The purpose of our research is not to mechanically emphasize the necessity of prevention and control policies, but to strive to reveal the decay and transmission mechanisms of RNA viruses. Perhaps the most important application value of this research lies in inspiring future vaccine development. Although we did not specifically find the specific nucleic acid sequence of this semi-infectious COVID-19 virus, we have macroscopically proven that this semi-infectious particle can cause a large number of asymptomatic infections in population infection scenarios. Therefore, future vaccine development should try to use these weakly pathogenic virus particles. The concept of using self-replicating live viruses as vaccines has been proposed and practiced by many people, such as the Peiyong Shi’s group [33], which used a partially genome-deleted COVID-19 virus as a vaccine. This self-replicating virus has many advantages. First, compared to traditional vaccines, its structure is infinitely closer to real viruses. Second, because of its weak replication ability and infectious attribute, this vaccine can stimulate antibodies production at extremely low injection doses, which helps to immunize large populations in a short period of time. Third, although the production of DVGs can lead to recombination, overall, due to the loss of template, the deletion is irreversible, and these deleted virus genomes only produce weaker viruses with more severe deletion. Therefore, the vaccine has good safety. What we need to do is to find such a defective virus sequence. This defective virus cannot be as severe as DVG, must have some self-replication ability, can induce the body to produce sufficient antibody concentrations, and the defect cannot be too weak, ensuring that all recipients are asymptomatic infections.

## 5 Conclusion

We conducted a systematic analysis of SARS-CoV-2 second-generation sequencing data using bioinformatics methods. The uniformity of the sequencing data revealed a higher level of genetic polymorphism in SARS-CoV-2 compared to DNA phages. This indicates a significant level of heterogeneity within the SARS-CoV-2 population, which may be attributed to mechanisms such as UTR region deletions, fragment breaks, or the formation of defective viral particles.

Further analysis of third-generation sequencing data provided insights into the replication process of SARS-CoV-2. Our findings suggest the presence of a replication element with low fidelity during viral replication, allowing for rapid base extension. However, the accuracy of the replication process is maintained through the corrective function of other replication elements at later stages. In terms of the genomic assembly, the Omicron strain demonstrated a significantly higher level of completeness compared to the Delta and Alpha strains. This disparity in genome assembly can effectively explain the heightened transmissibility of the Omicron variant relative to the Delta and Alpha variants.

By constructing a mathematical model to elucidate the process of viral polymorphism during infection, we can quantitatively assess the impact of viral heterogeneity on individual susceptibility. Our model provides insights into the beneficial role of defective viruses in host immune responses and elucidates their increasing proportions during the course of infection, which corresponds to changes in the transmissibility of infected individuals. Furthermore, by employing quantitative simulations of host-virus interactions, our model further clarifies why the virus is able to maintain its original genotype or length during evolution without rapidly deteriorating into low-virulence or non-virulent defective viruses.

Through the application of population infection models built upon mathematical frameworks, we can further examine the influence of climate conditions and social control measures on the manifestation of disease symptoms within populations. Our model effectively highlights the strong correlation between individual infection characteristics and the viral inoculum dose. Specifically, colder climate conditions and less stringent control measures can exacerbate the severity of symptoms within populations, thereby providing an explanation for the pronounced regional disparities in mortality rates observed during the course of COVID-19 infection.

Finally, our theoretical research also lays a foundation for future endeavors pertaining to the development of vaccines based on defective viruses. It is important to acknowledge the inherent uncertainties associated with our model, and its refinement will necessitate ongoing validation through additional experimental investigations and clinical data.

## Data Availability

Matlab codes can be accessed at:
https://github.com/zhaobinxu23/DVG_modeling

https://github.com/zhaobinxu23/DVG_modeling

## Code availability

Matlab codes can be accessed at: https://github.com/zhaobinxu23/DVG_modeling

## Author Contributions

Conceptualization, Z.X.; methodology, Z.X. and S.J.; writing—original draft preparation, Z.X. and Q.Z.; writing—review and editing, D.W. H.Z. and J.D.; funding acquisition, Z.X. and D.W. All authors have read and agreed to the published version of the manuscript.

## Acknowledgments

Dong-Qing Wei is supported by grants from the National Science Foundation of China (Grant No. 32070662, 61832019, 32030063), and Joint Research Funds for Medical and Engineering and Scientific Research at Shanghai Jiao Tong University (YG2021ZD02). The computations were partially performed at the Pengcheng Lab. and the Center for High-Performance Computing, Shanghai Jiao Tong University.

